# Genomics of severe and treatment-resistant obsessive-compulsive disorder treated with deep brain stimulation: a preliminary investigation

**DOI:** 10.1101/2023.04.15.23288623

**Authors:** Long Long Chen, Matilda Naesström, Matthew Halvorsen, Anders Fytagoridis, David Mataix-Cols, Christian Rück, James J. Crowley, Diana Pascal

**Author notes:** Correspond with Dr. Long Long Chen, M48, Karolinska Universitetssjukhuset Huddinge, 14186 Stockholm. These authors jointly supervised this work (James J. Crowley, Diana Pascal).

## Abstract

Individuals with severe and treatment-resistant obsessive-compulsive disorder (trOCD) represent a small but severely disabled group of patients. Since trOCD cases eligible for deep brain stimulation (DBS) probably comprise the most severe end of the OCD spectrum, we hypothesize that they may be more likely to have a strong genetic contribution to their disorder. Therefore, while the worldwide population of DBS-treated cases may be small (∼300), screening these individuals with modern genomic methods may accelerate gene discovery in OCD. As such, we have begun to collect DNA from trOCD cases who qualify for DBS, and here we report results from whole exome sequencing and microarray genotyping of our first five cases. All participants had previously received DBS in the bed nucleus of stria terminalis (BNST), with two patients responding to the surgery and one showing a partial response. Our analyses focused on gene-disruptive rare variants (GDRVs; rare, predicted-deleterious single-nucleotide variants or copy number variants overlapping protein-coding genes). Three of the five cases carried a GDRV, including a missense variant in the ion transporter domain of *KCNB1*, a deletion at 15q11.2, and a duplication at 15q26.1. The *KCNB1* variant (hg19 chr20-47991077-C-T, NM_004975.3:c.1020G>A, p.Met340Ile) causes substitution of methionine for isoleucine in the trans-membrane region of neuronal potassium voltage-gated ion channel KV2.1. This *KCNB1* substitution (Met340Ile) is located in a highly constrained region of the protein where other rare missense variants have previously been associated with neurodevelopmental disorders. The patient carrying the Met340Ile variant responded to DBS, which suggests that genetic factors could potentially be predictors of treatment response in DBS for OCD. In sum, we have established a protocol for recruiting and genomically characterizing trOCD cases. Preliminary results suggest that this will be an informative strategy for finding risk genes in OCD.

## INTRODUCTION

Obsessive-compulsive disorder (OCD) is a debilitating disorder with a lifetime prevalence of approximately 1-2% in the general population [1]. OCD often begins in adolescence and usually takes a chronic course if left untreated [2]. It can have substantial negative impacts on educational achievement, work productivity, and interpersonal relationships [3]. Moreover, it poses a significant burden and distress to caregivers [4], and OCD patients have a ∼10-fold higher risk of dying by suicide compared to those without an OCD diagnosis [5].

Approximately 10% of patients with OCD develop severe symptoms and do not respond to adequate treatment [6]. Treatment-resistant OCD (trOCD) patients typically have severe symptoms (e.g., a score >30 on the Yale-Brown Obsessive Compulsive Scale, Y-BOCS), are typically disabled for much of their life and do not gain symptom relief from conventional pharmacological or psychotherapeutic treatments [7]. Deep brain stimulation (DBS) has been an option for patients with chronically severe, disabling, and treatment-resistant OCD since 1999 [8], and to date, approximately 350 patients have undergone this procedure [9]. In DBS, thin electrodes connected to a neuro-pacemaker are introduced into subcortical central structures of the brain to modulate pathological neuronal activity with electrical current.

DBS is a routine neurosurgical treatment for Parkinson’s disease, dystonia, and tremor [10]. Evidence-based guidelines by the Congress of Neurological Surgeons recommend DBS for patients with severe and treatment-resistant OCD over the best medical treatment (level I and II) [11]. It has the advantage of being reversible, safe and the stimulation parameters can be adjusted to optimize treatment outcome. For OCD, several blinded randomized studies have demonstrated DBS to be effective in trOCD [12–16], with ∼60% of patients responding to treatment, defined as a ≥ 35% decrease in the Y-BOCS score [9]. The degree of improvement in OCD symptoms varies greatly, however, and the reason for this variability remains unknown. Therefore, identifying predictors of response to DBS in OCD, such as genetic markers, could facilitate the selection of patients for this invasive procedure.

Genetic factors account for ∼50% of the variance in risk for OCD, based on twin and family studies [17, 18], and first-degree relatives of affected individuals have ∼4-8 fold higher risk of developing OCD [17]. Regarding common variation, the first well-powered genome-wide association study (GWAS) of OCD was assembled by the Psychiatric Genomics Consortium (PGC) OCD Working Group and posted as a preprint in October 2021 [19]. This study was limited to European ancestry individuals (N = 14,140 OCD cases and N = 562,117 controls), but it did identify the first genome-wide significant locus for OCD. The PGC OCD Working Group is currently working on publishing a much larger OCD GWAS with >50,000 cases and >1 million controls. This marked increase in sample size has led to a major step forward in OCD genomics, in the form of ∼30 genome-wide significant loci where common genetic variation is associated with OCD (results presented at the 2022 World Congress of Psychiatric Genetics, Florence, Italy).

Regarding rare variation in OCD, two copy number variant (CNV) studies have shown that cases carry an increased burden of large deletions previously associated with neurodevelopmental disorders [20, 21], and larger studies are underway. Rare variants have also been implicated in OCD through two trio whole exome sequencing (WES) studies. Cappi et al [22] found that *de novo* mutations predicted to damage gene function are enriched in OCD probands (rate ratio, 1.52; *P* = 0.0005) and identified two high-confidence risk genes, each containing two damaging mutations in unrelated probands: *CHD8* and *SCUBE1*. The second study, from Halvorsen et al [23], reported WES results from the largest OCD cohort to date (1,313 OCD cases, consisting of 587 trios, 644 singletons, and 41 quartets). Compared to healthy controls, OCD cases carried an excess of loss of function (LoF) variants in genes intolerant to LoF variation. This was also true of *de novo* variants found in trios. In case-control analyses, the most significant single-gene result was *SLITRK5* (odds ratio = 8.8, P = 2.3 × 10^−6^), which is known to influence synapse formation.

In summary, OCD is a complex polygenic trait, and sequencing larger cohorts is needed to power the discovery of novel risk genes and provide greater insight into disease biology. We hypothesize that sequencing trOCD patients, in particular, may accelerate this process. Therefore, we have initiated a collection of DNA and clinical data from trOCD cases who qualify for DBS, and here we report results from WES and microarray genotyping of our first five cases.

## METHODS

### Participants

Participants (n = 5) were enrolled from two Swedish centers as part of two multi-center studies of DBS for OCD [24, 25]. All participants were of European ancestry and provided written informed consent. This study received ethical approval from the Swedish ethical review authority on 2014-11-26 (D. nr. 2014/1897-31).

### Inclusion criteria

Patients aged 18-65 years with trOCD, who have received DBS treatment, were eligible for this study. As is the convention in the field [26], trOCD cases had to meet each of the following criteria: OCD duration >5 years; Y-BOCS total score >30; three or more documented serotonin reuptake inhibitor (SRI) trials, including clomipramine (at least 10-12 weeks at an adequate dose); SRI augmentation for >4 weeks with at least one antipsychotic medication; an adequate trial of exposure and response prevention therapy (ERP) (intolerance or >15 sessions).

### Assessment

Phenotype data regarding demographic variables, previous treatments, and DBS stimulation parameters were collected for each patient and recorded by the responsible researcher at each site. Clinical assessments, including Y-BOCS [27], Montgomery-Åsberg Depression Rating Scale (MADRS) [28], Clinical Global Impression (CGI) [29], and the Euro-Qol 5-Dimensions Visual Analogue Scale (EQ5-D-VAS) [30] were administered before DBS and at one-year follow-up. Treatment response was defined as a 35% or greater decrease in the Y-BOCS score, while a 25-35% decrease was categorized as a partial response [31].

### Deep brain stimulation (DBS)

All participants were implanted with bilateral DBS systems from Medtronic PLC (Minneapolis, MN) in the Bed Nucleus of Stria Terminalis (BNST) under general anesthesia, using the Leksell frame model G [25]. For details of the surgical procedure, see published studies on the clinical effects and safety of BNST DBS in OCD [24, 25].

### Saliva sampling for DNA

Saliva samples were collected from participants using Oragene OG-500 kits from DNA Genotek, Inc (Ottawa, Canada). Genomic DNA was extracted and stored at the Karolinska Institutet Biobank at –80°C.

### Whole exome sequencing (WES) and read alignment

WES libraries were prepared from 50ng DNA using the Twist Human Core Exome sample preparation kit and probe panel (Twist Bioscience, #101897/101919/100578) with unique dual indexes (catalog #101308/09/10/11). The library preparation was performed according to the manufacturer’s instructions (DOC-001085 Rev. 1.0). Library quality was evaluated using an Agilent TapeStation and the D1000 ScreenTape Assay. The adapter-ligated fragments were quantified by qPCR using a library quantification kit for Illumina (KAPA Biosystems, Inc.) on a CFX384 Touch instrument (Bio-Rad Laboratories, Inc.) before cluster generation and sequencing.

Library preparation and sequencing were performed by the SNP&SEQ Technology Platform, a national unit within the National Genomics Infrastructure (NGI), hosted by the Science for Life Laboratory in Uppsala, Sweden. Sequencing was performed on an Illumina NovaSeq 6000 instrument (NSCS v 1.6.0/ RTA v 3.4.4). Demultiplexing and conversion to FASTQ format were performed using the bcl2fastq2 (2.20.0.422) software provided by Illumina.

WES read alignment was carried out according to GATK best practices [32]. Firstly, reads were aligned to the human reference genome hs37d5 using BWA v0.7.17 [33, 34]. Next, the BAMs were sorted and merged using Picard v2.2.4. The subsequent BAMs were indexed with samtools v1.8 [35]. Indel realignment on sample-level data in GATK v3.7 was conducted before read alignment was subjected to base recalibration. Lastly, we used verifyBamID v1.1.3 to determine if there was any evidence of contamination for any individual sample [36].

### Screening for gene-disruptive rare variants (GDRVs)

Single nucleotide variants (SNVs) and insertion/deletions (indels) were screened using the VCF-screen ‘gt’ function (https://github.com/Halvee/VCFScreen). For SNVs and indels, we required variants to be < 0.1% MAF in all gnomAD WES and WGS subpopulations. Furthermore, we required that variants had at least passing QC metrics, including PHRED-scaled likelihood (homozygous ref) ≥ 20, QD ≥ 2, MQ ≥ 40, sample coverage ≥ 10, and sample QUAL ≥ 20. All copy number variant (CNV) calls were made using XHMM 1.0 [37].

We specifically focused on SNVs, indels, and CNVs found in only 1 case sample. We defined a variant as ‘gene-disruptive’ if it met one of the following criteria: 1) SNV or indel that is protein-truncating (stop-gain, splice donor/acceptor disrupting, or frameshift annotation) within a protein-coding gene; 2) missense SNV with Missense badness, PolyPhen-2, and Constraint (MPC) > 3; 3) CNV deletion or duplication impacting at least one protein-coding base. We only considered the subset of GDRVs that impacted a ‘constrained’ gene, defined as having gnomAD v2.1.1 pLI > 0.9 (n=3063 genes total) as these genes have a substantial depletion of GDRVs in the general population [38, 39].

### Array-based genotyping and CNV calling

All samples were genotyped using the Illumina Global Screening Array version 3 at LIFE&BRAIN GmbH in Bonn, Germany. We ensured sample concordance by verifying genotypes at 24,218 sites where both exome and post-imputation genotype array data were available [19]. We extracted all CNV calls from a large case/control OCD study (manuscript in preparation) where these samples were included and verified that the 15q11.2 deletion and the 15q26.1 duplication were called from these array data as well.

## RESULTS

### Socio-demographic characteristics

***Table 1*** presents the socio-demographic characteristics of our cohort. The mean duration of illness was 20.6 years, and the range of ages when participants received DBS treatment was 27 to 53 years old. All participants were of European ancestry, and we enrolled three males and two females. Three of the five patients had comorbid ADHD, while none had a history of tics or Tourette syndrome. Just one of the five participants attained a university degree, underscoring the negative impact of trOCD on educational achievement. Four of the five patients self-reported at least one first-degree relative with a diagnosis of OCD.

**Table 1.** Participants’ demographic variables. Only essential details are given, in order to minimize the risk of identifiability.

| Patient number | Duration of illness (years) | Comorbidities | OCD in first-degree relative? | Highest Education |
| --- | --- | --- | --- | --- |
| 1 | 5 | ADHD | Yes | High school |
| 2 | 46 |  | Yes | High school |
| 3 | 11 | ADHD, ASD | Yes | University |
| 4 | 16 | ADHD | Yes | Elementary school |
| 5 | 25 |  | No | High school |
| Autism spectrum disorder (ASD), attention deficit hyperactivity disorder (ADHD), female (F), male (M). |  |  |  |  |

### DBS treatment outcome at one-year follow-up

All participants received bilateral DBS in the BNST, as previously reported by Naesström et al. and Menchón et al. [24, 25]. Stimulation voltages ranged from 3.2 to 5.5 Volts, pulse width ranged from 90 to 210 milliseconds, and all patients had pulses delivered at a frequency of 130 Hertz (see ***Table 2***). Adverse events were mild, rare, and transient (i.e., acute hypomania and fatigue). Changes in clinical scales are presented in ***Table 3***. Patients #1 and #2 were responders, and patient number #3 was a partial responder. Patient #5 ran out of batteries at the one-year follow-up but was a responder after the battery change.

**Table 2.** Deep brain stimulation parameters at one-year follow-up.

| Patient number | DBS location | Voltage (Volts) | Frequency (Hertz) | Pulse width (msec) | Adverse events |
| --- | --- | --- | --- | --- | --- |
| 1 | BNST | 3.2 | 130 | 90 | Hypomania |
| 2 | BNST | 5.0 | 130 | 120 | Fatigue |
| 3 | BNST | 4.9 | 130 | 90 |  |
| 4 | BNST | 3.8 | 130 | 90 |  |
| 5 | BNST | 5.5 | 130 | 210 |  |
| Bed nucleus of stria terminalis (BNST). |  |  |  |  |  |

**Table 3.** Clinical assessment before and at one-year follow-up.

| Patient number | Y-BOCS before | Y-BOCS after | MADRS before | MADRS after | EQ5D-VAS before | EQ5D-VAS after | CGI-S before | CGI-S after | Response status |
| --- | --- | --- | --- | --- | --- | --- | --- | --- | --- |
| 1 | 30 | 14 | 34 | 9 | 15 | 55 | 7 | 5 | Responder |
| 2 | 35 | 14 | 31 | 18 | 10 | 50 | 6 | 4 | Responder |
| 3 | 35 | 23 | 37 | 32 | 48 | 45 | 6 | 5 | Partial responder |
| 4 | 31 | 24 | 26 | 23 | 70 | 80 | 5 | 5 | Non-responder |
| 5 | 37 | 33 | 9 | 12 | 27 | 30 | 7 | 7 | Non-responder |
| Yale-Brown Obsessive-Compulsive Scale (Y-BOCS), Montgomery-Åsberg Depression Rating Scale (MADRS), Clinical Global Impression (CGI), Euro-Qol 5-Dimensions (EQ5-D). |  |  |  |  |  |  |  |  |  |

### GDRVs detected

A total of three unique GDRVs in constrained genes were detected in three different individuals. Patient #2 had a missense variant in the ion transporter domain (S5) of the gene *KNCB1* (hg19 chr20-47991077-C-T, NM_004975.3:c.1020G>A, p.Met340Ile) (see ***Figure 1***).

**Figure 1.**
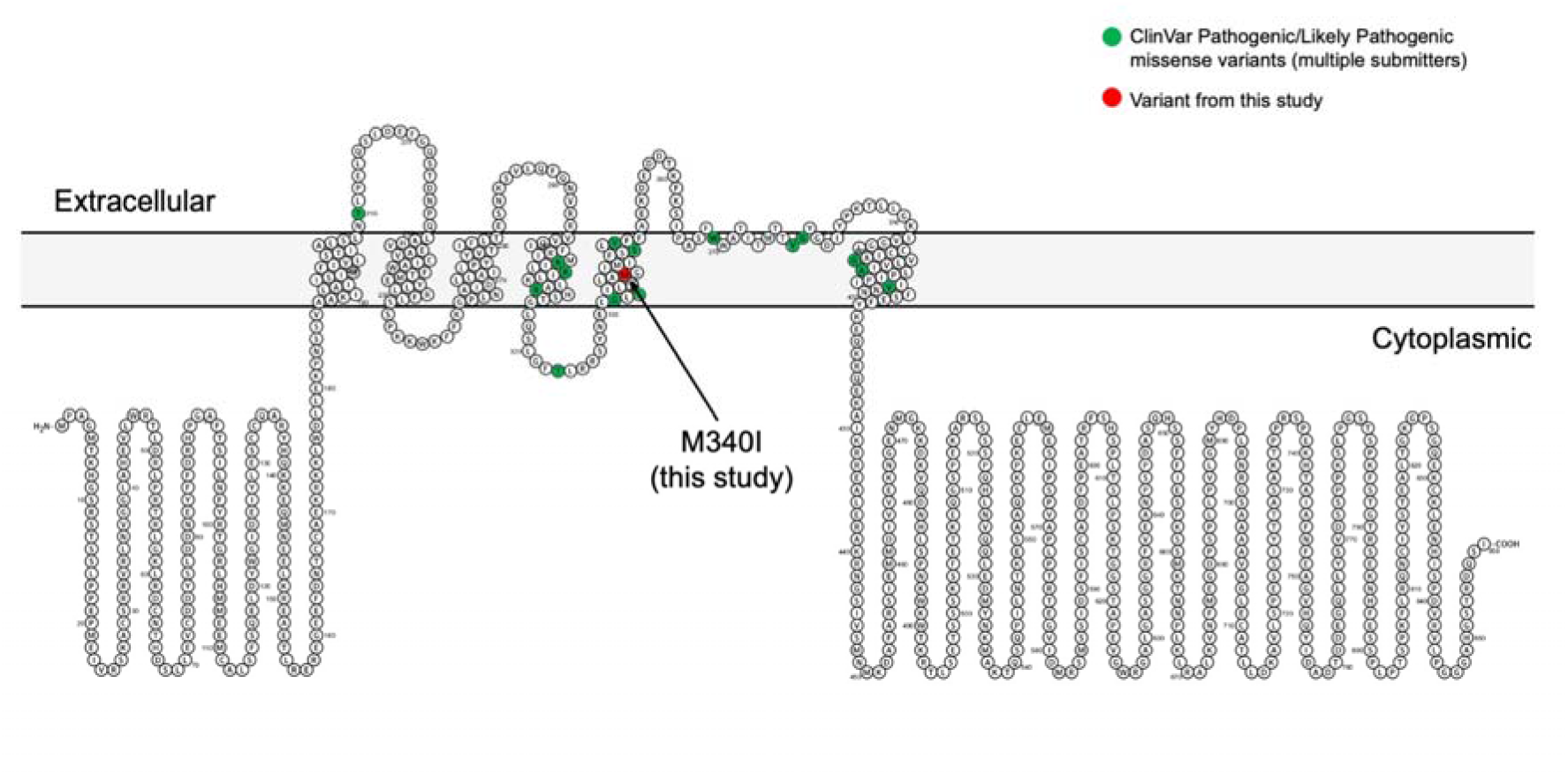
A *KCNB1* missense variant identified in trOCD case #2 colocalizes with known pathogenic missense variants. *KCNB1* encodes the potassium voltage-gated channel subfamily B member 1 and has six transmembrane domains. Shown in green are amino acids changed by missense variants classified as pathogenic or likely pathogenic in ClinVar. Shown in red is the variant identified in this study (hg19 chr20-47991077-C-T, NM_004975.3:c.1020G>A, p.Met340Ile). Like most of the pathogenic or likely pathogenic variants, M340I localizes to a transmembrane domain that helps form the pore through which potassium passes.

It is absent from gnomadAD v2.1.1, and the variant is in a region highly depleted from missense variants (constrained coding region (CCR) percentile of 99.8%) [40]. Moreover, this variant is predicted to be deleterious as it has an MPC (Missense badness, PolyPhen-2, and Constraint) score of 3.306, which is greater than 99.5% of all MPC annotations in MPC v2 [41]. Furthermore, as shown in ***Figure 1***, the *KCNB1* variant identified here clusters with other previously reported pathogenic/likely pathogenic missense variants in *KCNB1* (N = 15 high-confidence ClinVar variants supported by multiple submitters).

Patient #3 had a 768 kb deletion call in 15q11.2 that disrupted 4 genes, 2 with pLI > 0.9 (coordinates: hg19 chr15:22835916-23280000) (see ***Figure 2a***). The breakpoints of this deletion match those of previously reported recurrent CNVs in this region known to increase the risk of neurodevelopmental disorders (see discussion for details). Patient #4 had a 287 kb duplication in 15q26.1 (coordinates: hg19 chr15:89169441-89456550) (see ***Figure 2b***). This duplication disrupted five genes, one with pLI > 0.9 (*ACAN*). We note that while this gene has a high pLI, it has a lower probability of triplosensitivity (pTriplo = 0.293) [42]. We also conducted CNV calling in genotype array data, and were able to detect both CNVs, with both impacting the same core sets of genes indicated. No GDRVs were detected in patients #1 and #5.

**Figure 2.**
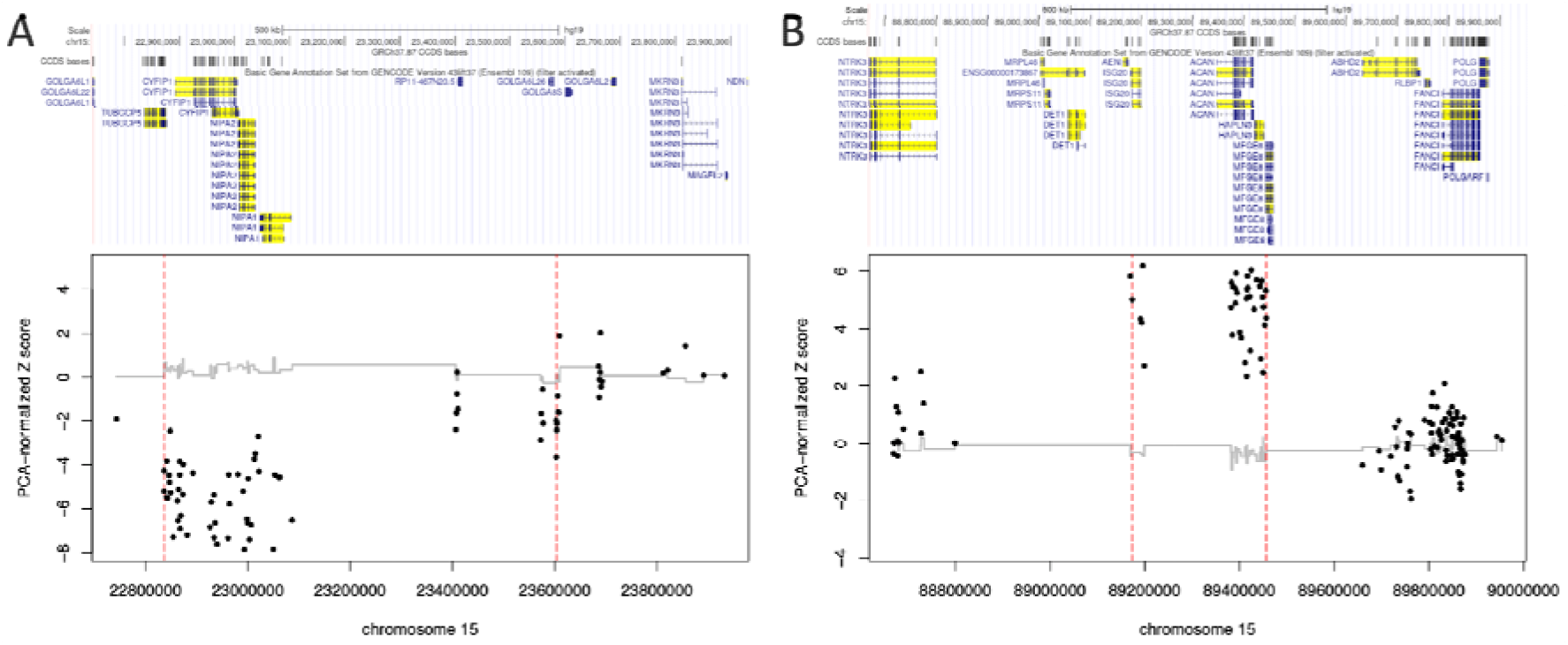
Gene-disruptive rare CNVs called in exome sequence data from trOCD cases #3 (A) and #4 (B). The red vertical dashes indicate the boundaries of the CNV call. Patient #3 has a previously described 768 kb neurodevelopmental deletion in 15q11.2 that disrupts 4 genes. Patient #4 has a novel 287 kb duplication in 15q26.1 that disrupts 5 genes.

## DISCUSSION

In this preliminary genomic investigation of trOCD, we found that three of the five individuals studied carried a GDRV. While a much larger sample is required to make definitive conclusions, the rate of GDRVs in trOCD may be higher than in more typical OCD. For example, in the largest OCD exome sequencing study to date [23], of 771 OCD trios meta-analyzed, 27 (3.5%) carried a *de novo* SNV or indel that could be classified as a GDRV impacting a gene with pLI > 0.9 (using the same criteria defined in this paper). It is conceivable that GDRVs could be enriched in trOCD cases and contribute to the severity of their OCD symptoms and treatment resistance.

The most compelling variant found in this study is patient #2’s missense mutation in *KCNB1* (Met340Ile). This gene encodes a voltage-gated potassium channel, KV2.1, an important regulator of neuronal excitability [43]. *KCNB1* is evolutionary conserved, highly expressed in the frontal cortex, and depleted of missense and protein-truncating variation in the general population. In fact, no protein-truncating variants in *KCNB1* are found in gnomAD (v2.1.1). There are, however, a number of rare, deleterious missense variants in *KNCB1* associated with developmental and epileptic encephalopathies [44]. These variants tend to be *de novo* in origin and cluster in the channel’s transmembrane domains, including the ion transporter domain (S5), which is where we observed Met340Ile in a trOCD patient (see ***Figure 1***).

Besides encephalopathies, recent reports have implicated *KCNB1* variants in a spectrum of phenotypes, including neurodevelopmental disorders [45, 46]. For example, patients with missense variants in neighboring amino acids (positions 334 and 347) reportedly suffer from autism spectrum disorder, intellectual disability, and/or epilepsy, but the severity of symptoms differs [46]. Interestingly, patient #2 responded to DBS treatment, suggesting that GDRVs are not necessarily negative predictors of DBS treatment response. This is consistent with a previous case report where compulsive behavior diminished in a patient with a 9q34.3 deletion after DBS treatment [47].

The clinical significance of the two CNVs (15q11.2 deletion and 15q26.1 duplication) are uncertain, according to the American College of Medical Genetics [48]. Duplication of 15q26.1 has not been associated with neurodevelopmental disorders previously, so the clinical significance is unknown. The 15q11.2 deletion, however, has been described as a neurodevelopmental CNV with a modest effect size. For example, a meta-analysis estimated the overall effect size of the deletion to be a 4.3-point decrease in IQ and a slightly increased risk of schizophrenia and epilepsy [49]. However, there is considerable phenotype variability among 15q11.2 deletion carriers in the literature [50, 51]. Multiple brain-expressed genes are located in this region of 15q11.2, most notably *CYFIP1*, which is thought to impact neuronal function and morphology [52–54].

The primary limitation of this study is the limited sample size. However, there are approximately 300 OCD cases in the world who have undergone DBS [26, 55] and additional cases who have had ablative surgeries. Increasing sample size through international collaboration in the future is crucial. Due to the sample size, statistical inference testing was not viable. In addition, based on single cases, we cannot conduct formal association tests between the genetic findings and OCD. Since we lack sequence data from parents, we are also unable to identify which variants are *de novo* in origin. This information is critical for formally classifying a variant (particularly those with missense annotation) as pathogenic or likely pathogenic. The high rate of comorbidity with ASD and ADHD suggests that these GDRVs may be associated with a cluster of neurodevelopmental disorders.

In conclusion, this is the first OCD exome sequencing study focused on patients with severe and treatment-resistant OCD. We demonstrate that small but well-defined subgroups of patients with psychiatric disorders are relevant for in-depth genetic analysis. The number of GDRVs uncovered in this cohort corroborates our hypothesis of associations between genetic load with symptom severity and prognosis of OCD.

## Data Availability

All data produced in the present study are available upon reasonable request to the authors.

## ACKNOWLEDGEMENTS

Sequencing was performed by the SNP&SEQ Technology Platform in Uppsala. The facility is part of the National Genomics Infrastructure (NGI) Sweden and Science for Life Laboratory. The SNP&SEQ Platform is also supported by the Swedish Research Council and the Knut and Alice Wallenberg Foundation. We are grateful for the contributions of our study participants. We thank Stephanie B. Crowley for assistance with the manuscript.

